# Multiple drug arrests reported to the Maine Diversion Alert Program more frequently involved carisoprodol, amitriptyline, or quetiapine

**DOI:** 10.1101/2021.05.25.21257786

**Authors:** Maaz Siddiqui, John P. Piserchio, Misha Patel, Jino Park, Michelle Foster, Clare E. Desrosiers, John Herbert, Kenneth L. McCall, Brian J. Piper

## Abstract

**Background:** Much of the blame of the increasing death toll by drug overdoses has justifiably been attributed to the United States’ current opioid epidemic. However, nearly 80% of overdoses related to opioids involve another drug substance or alcohol. The objective of this study was to elucidate overrepresentation of drugs in polypharmacy arrests by identifying drugs that were more likely to be found in conjunction with other substances, using the drug arrest data provided by the Maine Diversion Alert Program (DAP).

**Methods:** Single drug arrest and multiple drug arrest totals reported to the DAP were examined. Drugs involved in the arrests were classified by Drug Enforcement Administration Schedule (I-V or non-controlled prescription) and categorized into five drug families: hallucinogens, opioids, sedatives, stimulants, and miscellaneous. Multiple drug arrest totals were compared to single drug arrest totals to create a Multiple-to-Single Ratio (MSR) specific to each drug family and each drug. Chi-square approximations without Yates correction and two-tailed P values were used to determine statistical significance through GraphPad’s 2×2 contingency tables.

**Results:** Over three-fifths (63.8%) of all arrests involved a single drug. Opioids accounted for over-half (53.5%) of single arrests, followed by stimulants (27.7%) and hallucinogens (7.7%). Similarly, nearly two-fifths (39.6%) of multiple arrests were opioids, followed by stimulants (30.8%) and miscellaneous (13.0%). Miscellaneous family drugs were recorded with the highest Multiple-to-Single Ratio (1.51), followed by sedatives (1.09), stimulants (0.63), opioids (0.42), and hallucinogens (0.35). Carisoprodol (8.80), amitriptyline (6.34), and quetiapine (4.69) had the highest MSR values and therefore were the three most overrepresented drugs in polysubstance arrests.

**Conclusion:** The abuse of opioids, both alone and in conjunction with another drug, deserves continued surveillance in public health. In addition, common prescription drugs with lesser-known misuse potential, especially carisoprodol, amitriptyline, and quetiapine, require more attention by medical providers for their ability to enhance the effects of other drugs or to compensate for undesired drug effects.

## INTRODUCTION

Deaths attributed to drug overdose in the United States (US) have continued to rise over the last twenty years resulting in over 70,000 deaths in 2019 alone. Although much of the focus has rightfully been placed on the opioid drug class, deeper investigations into the patterns of drug misuse have exposed a multitude of other drug classes that contribute to overdose-related deaths, including stimulants such as cocaine and methamphetamine, depressants such as benzodiazepines, and even antidepressants (1). In fact, toxicology studies of all synthetic opioid-related overdose deaths in 2016 showed that nearly 80% of the death involved another drug or alcohol (2). Polysubstance drug use is becoming increasingly common in the US. An analysis of 2,244 opioid overdose-related deaths in Massachusetts showed that 36% of the deaths involved a stimulant and 46% involved a non-stimulant substance. Only 17% of the 2,244 deaths involved solely an opioid. In 2018, there were over 16,000 drug overdose deaths in 24 states and Washington DC, a third of which included both opioids and stimulants (3). Polysubstance misuse, abuse, and addiction continues to impact millions of American’s every day and new information is needed to develop tools to combat this pervasive public health crisis.

The Maine Diversion Alert Program (DAP) was an informational resource that facilitated communication about drug arrests between the criminal justice and healthcare fields (4–10). An annual DAP report made the counter-intuitive finding that non-controlled pharmaceuticals were more likely to be involved in arrests that involved other drugs (11). The objective of this study was to elucidate overrepresentation of drugs in polypharmacy arrests by identifying the specific drugs that were more likely to be found with other substances. The DAP has previously identified some drugs not commonly thought to have pronounced addictive profiles like gabapentin (12). Drug addiction is complex, and we hypothesized that a variety of agents, perhaps even those not commonly thought to contribute to drug addiction (e.g. amitriptyline, carisoprodol, quetiapine), would be found in combination with other more common illicit drugs like heroin and cocaine. This hypothesis was based on the premise that drug users experiment and discover ways to manipulate the pharmacodynamics or pharmacokinetics of their primary drug by using multiple drugs concurrently to enhance euphoria or hallucinogenic properties and control adverse effects. By investigating polysubstance arrest patterns, we hoped to contribute a greater understanding of drug addiction and the role polysubstance misuse plays in overdose-related mortality, but at an earlier juncture where corrective measures could be implemented.

## METHODS

### Subjects

Subjects included individuals reported to the Maine DAP for drug arrests (N = 9,216, 31.38% female; age = 32.98, SD = 9.85, min = 18, max = 83, a minority 246 (2.67%) of individuals did not have age information). Additional information concerning sex, town of residence, county, offense, drug(s), drug category, federal schedule, Maine drug schedule, number of drugs, date and year of charge, law enforcement agency, and agency category were submitted by local, county, state, and federal law enforcement personnel.

### Procedures

Drug arrest data was documented through the Maine DAP during its course of its statewide operation from June 2013 to early 2018. The DAP ceased operation in early 2018 due to insufficient funding. Drugs (N = 170) involved in the arrests were classified by Drug Enforcement Administration Schedule as I-V or non-controlled prescription in accordance with the US 1970 Controlled Substances Act and updates. Substances were then categorized into drug families as: hallucinogens (e.g., lysergic acid diethylamide (LSD) and methylenedioxymethamphetamine (MDMA)); opioids (e.g., morphine, methadone); sedatives (e.g., carisoprodol, zolpidem); stimulants (e.g., amphetamine, methylphenidate); and miscellaneous pharmaceuticals including substances not classified under the other drug families that are typically manufactured by pharmaceutical companies (e.g., amitriptyline, quetiapine). Additional information about the families may be found elsewhere (4-11). Unspecified or unknown drugs at the time of arrest (N = 1,108) or drug paraphernalia (N = 85) were excluded from further analysis. Procedures were approved by the Institutional Review Board of the Wright Center of the Graduate Medical Education of Scranton.

### Data Analysis

Summons and indictments were categorized for simplicity as “arrests”. Single (N = 6,441) and multiple (N = 3,660) drug arrests totals were compared. For the assessment of multiple drug arrests, each unique drug reported per offense was analyzed. Figures were created using GraphPad Prism, version 9.1.0. Multiple drug arrest totals were compared to single drug arrest totals to create a Multiple-to-Single Ratio (MSR) specific to each drug family and each drug. For example, a substance that accounted for 10% of multiple drug arrests and 5% of single drug arrests would have a MSR of (10%)/(5%) or 2.0. An MSR > 1.0 indicated overrepresentation and an MSR < 1.0 indicated underrepresentation. Agents that were involved in less than 10 multiple arrests were not reported in Figure 2 so that smaller values did not exert an undue impact on the MSR. Chi-square approximations without Yates’ correction and two-tailed p-values were used to determine statistical significance through GraphPad’s 2×2 contingency tables. An alpha of <0.05 was considered statistically significant.

## RESULTS

Multiple and single drug arrestees were similar demographically. Multiple (31.51% female) and single (31.35% female) were equivalent in terms of sex. Multiple (33.36 ± 9.56 years) and single (32.91 ± 9.91 years) did not differ appreciably based on average age.

Over three-fifths (63.8%) of all drug arrests involved a single drug. Opioids (e.g. heroin, oxycodone, and buprenorphine) accounted for over-half (53.5%) of single arrests, followed by stimulants (27.7%, e.g. cocaine, crack, and methamphetamine) and hallucinogens (7.7%, e.g. marijuana, bath salts, and MDMA). Specifically, the five most common drugs in single drug arrests were heroin (27.15%), oxycodone (10.48%), cocaine (10.36%), buprenorphine (9.05%), and (5) crack (7.72%).

Similar to the single drug arrest data, nearly two-fifths (39.6%) of multiple arrests were opioids, followed by stimulants (30.8%) and miscellaneous (13.0%). Likewise, the five most common drugs in multiple drug arrests were heroin (17.05%), cocaine (11.37%), crack (9.70%), buprenorphine (8.17%), and (5) oxycodone (5.63%).

Figure 1 shows the overall MSR of each drug family. The drug families that had the highest overall MSR values were miscellaneous (1.51) and sedatives (1.09) which were significantly over-represented for arrests. Stimulants (0.63), opioids (0.42) and hallucinogens (0.35) were significantly under-represented for arrests. Individual agents that were found in both single and multiple drug arrests (N = 34) were explored further.

**Figure 1.**
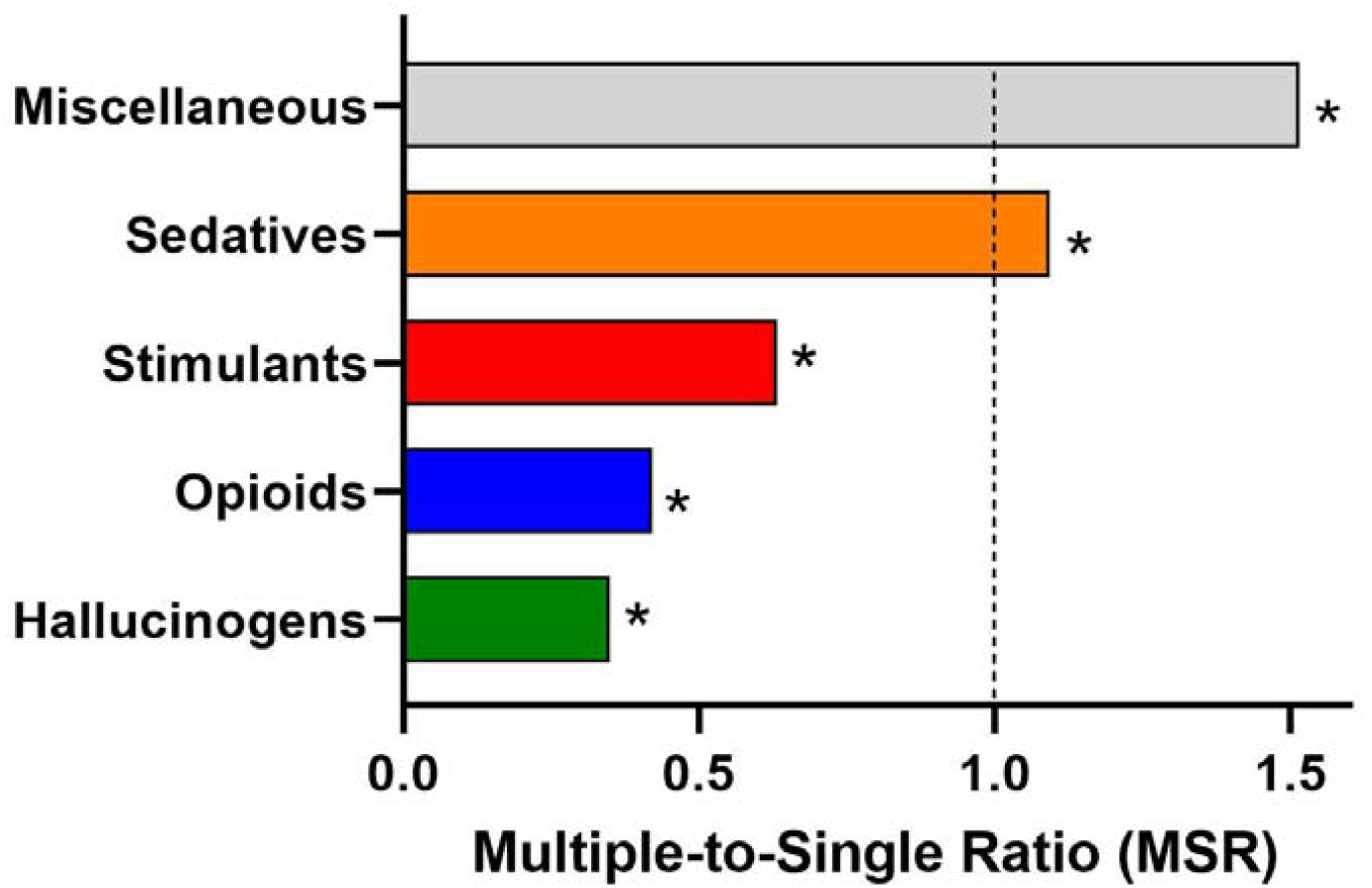
Multiple-to-Single Ratio by drug families for arrests reported to the Maine Diversion Alert Program. *p < 0.05 versus all other families.

Figure 2 shows the MSR for each of the 34 drugs involved in both single and multiple drug arrests. Sedatives had the highest MSR, followed by miscellaneous, opioids, stimulants, and hallucinogens. Firstly, carisoprodol (8.80), zolpidem (2.93), and diazepam (2.86) had the three highest MSRs within the sedative drug family, while alprazolam (1.38) was the lowest. Secondly, amitriptyline (6.34), quetiapine (4.69), and clonidine (2.84) had the three highest MSRs within the miscellaneous drug family, while bupropion (1.48) was the lowest. Thirdly, morphine (3.06), methadone (1.64), and fentanyl (1.35) had the three highest MSRs within the opioid drug family, while oxycodone (0.54) was the lowest. Fourthly, amphetamine (2.27), methylphenidate (1.97), and lisdexamfetamine (1.29) had the three highest MSRs within the stimulant drug family, while methamphetamine (0.44) was the lowest. Lastly, LSD (1.94), MDMA/MDA (1.16), and hashish (0.71) had the three highest MSRs within the hallucinogen drug family, while α-PVP/bath salt (0.34) was the lowest.

**Figure 2.**
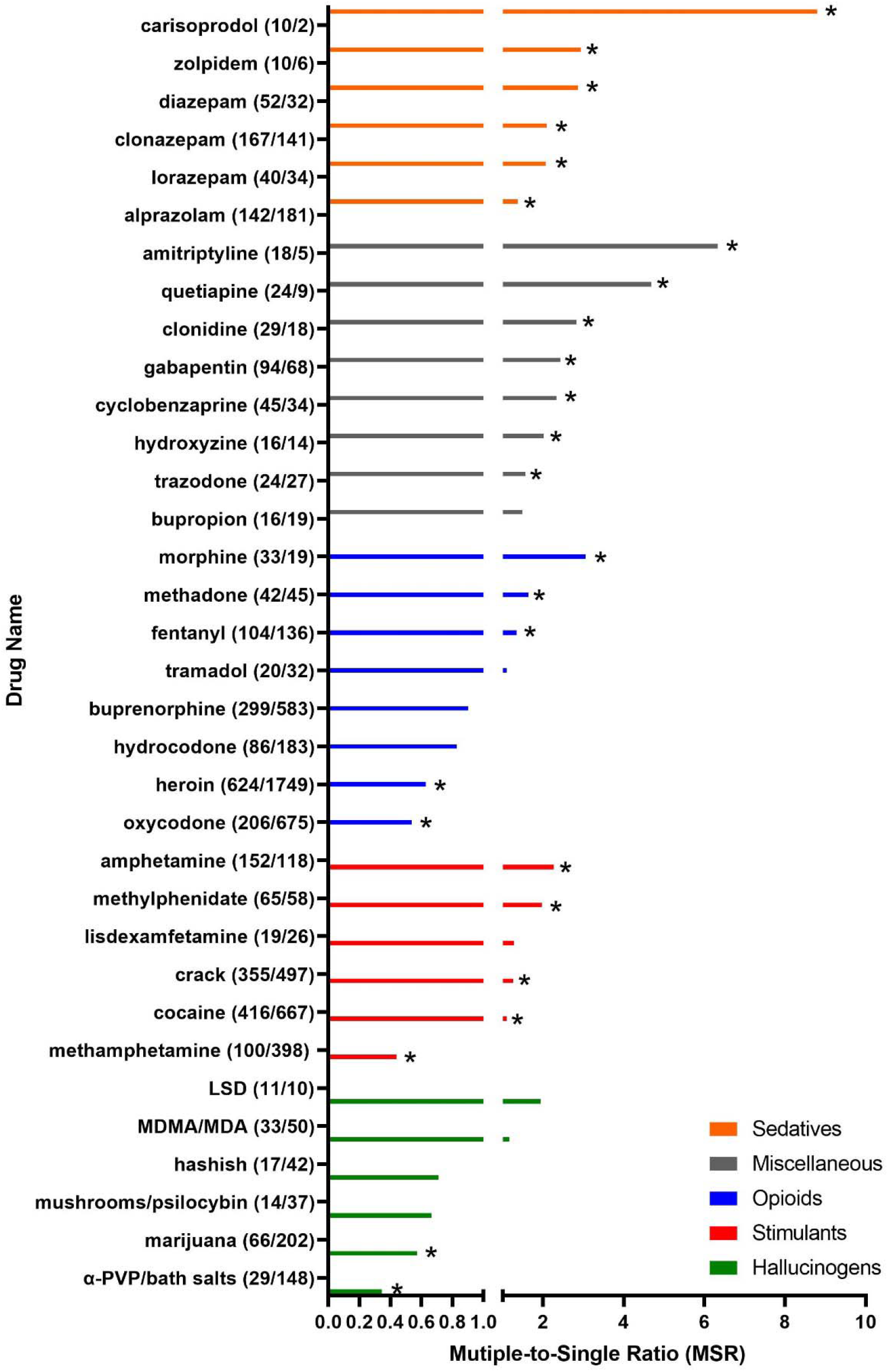
Multiple-to-Single Ratio for specific drugs group by drug families as reported by the Maine Diversion Alert Program. Each drug’s multiple and single drug arrests are in parentheses. *p < 0.05

## DISCUSSION

We examined drugs that were overrepresented in multiple drug arrests relative to single drug arrests as reported by the Maine DAP. The substances with the highest, and significant, overrepresentation (MSR > 1.0) in our analysis were carisoprodol (8.80), amitriptyline (6.34), and quetiapine (4.69). Polysubstance misuse in the United States is commonly associated with notable drug combination patterns, such as the concurrent use of stimulants with opioids to create a more tolerable adverse effect profile of each drug. Illicitly manufactured drugs such as heroin, fentanyl, cocaine, and methamphetamine are known to have a large impact on polysubstance overdose-related mortality (13), however, our understanding of how prescription drugs impact polysubstance drug addiction is still poorly understood.

Interestingly, the drugs that were most under-represented in our analysis (MSR < 1.0) were methamphetamine, heroin, bath salts, oxycodone, and marijuana. While drugs with a higher index of abuse and comorbidity such as heroin and methamphetamine are well known to result in morbidity and mortality, our data suggests that there are other commonly prescribed drugs that may be more pervasive in the development of polysubstance drug addiction and subsequent enhanced drug misuse behavior. What follows is a discussion that reviews the pharmacology of carisoprodol, amitriptyline, and quetiapine, and current theories of how each drug may contribute to polysubstance drug addiction.

The tricyclic antidepressant (TCA) amitriptyline also has analgesic properties that lead to its use in treating depression, neuropathic pain, migraines, and fibromyalgia (14). TCAs exert their effect by inhibiting the reuptake of norepinephrine and serotonin in adrenergic and serotonergic neurons. As one of the first antidepressants to be discovered, TCAs have more recently fallen out of favor in treating depression due to their antimuscarinic and cardiac adverse effect profile. However, they remain one of the more commonly prescribed drugs for their efficacy in treating pain that is unresponsive to narcotics. Despite a lack of addictive chemical components, amitriptyline’s sedative and anti-anxiolytic effects have led to misuse of the drug. Case reports of amitriptyline abuse describe feelings of “relaxation, giddiness, and contentment” as well as “a buzz… numbed up… calming” effects in patients who were found to have elevated levels of amitriptyline and no other substance in the emergency department (15,16). Dosages of 100-200 mg/day are sufficient to cause euphoric effects and potentiate abuse (17). Further, due to the nature of the diseases that TCAs treat, many patients who use amitriptyline have other comorbid conditions such as diabetes that necessitate polysubstance treatment or psychiatric afflictions that foster higher risk for mixed-drug abuse. Although amitriptyline is not listed as a controlled substance by the US Drug Enforcement Administration, it is frequently one of the drugs involved in suicidal and accidental multi-drug overdoses (18). Interestingly, among 346 people enrolled in a methadone maintenance program, 25% admitted to taking amitriptyline in order to achieve feelings of euphoria, not only indicating that amitriptyline misuse is not uncommon, but also suggesting that it may have synergistic effects with opioids (19). Altogether, this data warrants further investigation as to whether individuals were intentionally co-administering amitriptyline to enhance the sedating effects of opioids or benzodiazepines.

The muscle relaxer carisoprodol is indicated for use for acute musculoskeletal pain with guidelines suggesting a maximum prescription length of two to three weeks. Withdrawal is a concern with long-term or high-dose users. In the last two decades, carisoprodol use and abuse increased to the point of requiring scheduling as a controlled substance in January 2012. The exact mechanism of action is not known; however, research suggests that it causes a depressant effect at GABA receptors. This effect is often augmented with opioids, benzodiazepines, or alcohol. A 2012 national survey found that millions of individuals report using carisoprodol for non-medical reasons (20). Between 2011-2016, carisoprodol ranked amongst the top 15 drugs involved in drug overdose deaths. Despite the increased protections surrounding this drug, higher vigilance and judicious prescriptions from clinicians will benefit patients.

The second-generation antipsychotic quetiapine has a diverse mechanism of action. Quetiapine shows greater affinity for the histamine (H_1_), adrenergic (α1), muscarinic (M_1/3_) and serotonergic (5-HT_2A/C_) receptors as well as the norepinephrine transporter relative to the dopamine (D_2_) receptor (21). Quetiapine is normally prescribed to treat psychotic and mood conditions, including schizophrenia and bipolar disorder. There is a growing concern that quetiapine is being commonly prescribed and used as off-label prescriptions for dementia, insomnia, anxiety, and po st-traumatic stress disorder (PTSD) and may be contributing to adverse outcomes in vulnerable populations, such as those with mental health illnesses (22). Quetiapine is also the most documented antipsychotic bought and sold illicitly which is most likely due to the drug’s relatively low risk for the development of dystonia and extrapyramidal symptoms due to its low affinity for D_2_ receptors when compared to other antipsychotics. Due to quetiapine’s growing use with other drugs, its role in drug abuse and polysubstance use has been investigated. Quetiapine has unique sedative and anxiolytic effects due to its antagonistic effect on H_1_ receptors and serotonin receptors, respectively. This drug effect profile has resulted in quetiapine’s frequent use with stimulants such as amphetamines and cocaine to reduce the feelings of anxiety that come with using stimulants (23). By controlling the adverse effects associated with stimulant use, we suspect this can lead to higher drug doses and higher risk of mortality. Additionally, studies on rodents have shown that administration of antihistamines increases the release of dopamine in the ventral striatum similar to other addictive psychoactive substances such as cocaine, and thus, quetiapine may also have reward properties that contribute to drug misuse and addiction. Alternatively, quetiapine has been often found in forensic autopsies and is thought to be used in combination with opioids to reinforce the sedative effects of these drugs (24).

Our evidence suggests that polysubstance drug use is characterized by a pattern of drug use that works to enhance the effects of other drugs (carisoprodol and amitriptyline) or to compensate for undesired drug effects (quetiapine). While much attention in polysubstance literature focuses on the morbidity and mortality related to scheduled drug use, such as heroin and cocaine, it is important to understand how polysubstance use may develop through the use of commonly prescribed drugs. In our investigation of DAP’s database of individuals arrested and charged with possession of drugs, specific drugs such as carisoprodol, amitriptyline, and quetiapine were each found to be most likely used with other drugs. The basic pharmacology and current understanding of each drug’s addictive potential indicates that these drugs can contribute to polysubstance drug addiction and a pattern of drug use that encourages the use of more potent and riskier drugs.

In addition to pharmacological factors, legal practices and the broader sociocultural environment also contribute to the pattern of substances which were over, and under, represented in multiple drug arrests. Data was collected between 2013 and 2018. The largest city in the state, Portland, decriminalized marijuana in 2013 and a statewide referendum legalizing recreational marijuana was passed in 2016. Law enforcement officers have some discretion in making arrests. This descriptive dataset cannot distinguish whether some individuals found in the possession of only a single non-scheduled prescription drug were not arrested or whether charges for carisoprodol, amitriptyline, or quetiapine were more likely to be issued when “harder” illicit substances were also discovered by officers.

In conclusion, when analyzing multi-drug arrests reported by Maine’s Diversion Alert Program (DAP) between 2013 and 2018, we observed an overrepresentation of commonly prescribed drugs such as amitriptyline, carisoprodol, and quetiapine and an underrepresentation of drugs with a higher index of abuse such as methamphetamine and heroin. These findings, in addition to amitriptyline, carisoprodol, and quetiapine’s pharmacological plausibility of manipulating illicit drug use, supports the notion that polysubstance use with commonly prescribed pharmaceutical medications may contribute to riskier illicit drug use tendencies. Further investigation is warranted to better understand the role polysubstance use plays in the development of drug addiction, specifically with commonly prescribed drugs, and how healthcare professionals can identify these patterns in clinical practice to reduce drug misuse morbidity and mortality.

## Data Availability

Per the Data Use Agreement signed with the Diversion Alert Program, this information is not publically available.

## Acknowledgements

The contributions of Stephanie D. Nichols, PharmD, Kevin J. Simpson, MD, Matthew T. Moran, MD, and Dipam T. Shah, MD are gratefully acknowledged.

